# Prevalence of Microvascular Complications and Associated Risk Factors among Diabetes Mellitus Patients attending Nyeri County Referral Hospital, Kenya: a cross-sectional Study

**DOI:** 10.1101/2024.02.15.24302876

**Authors:** Rachael Ireri, Gideon Kikuvi, Susan Mambo, Besty Cheriro

**Author notes:** Corresponding Author: Rachael Ireri (.

## Abstract

Microvascular complications are one of the major causes of morbidity and mortality worldwide among patients with diabetes mellitus (DM). In Kenya, Nyeri County has the highest prevalence of diabetes mellitus compared to other counties. More than 50% of Nyeri County Referral Hospital (NCRH) admissions result from non-communicable diseases (NCDs) and over 55% of hospital deaths are attributable to NCDs. This study therefore sought to assess the prevalence of microvascular complications and the associated risk factors among patients attending Nyeri County Referral Hospital in Kenya.

A hospital-based cross-sectional study was conducted on 314 DM patients on follow-up at NCRH from August 2022 to October 2022. The socio-demographic, anthropometric, biochemical, and clinical data were recorded using a structured survey. All the recruited patients underwent an extensive examination for the presence of Microvascular complications like neuropathy, retinopathy, and nephropathy. Data were analyzed using STATA version 17. Univariate and multivariate logistic regression analyses are used to determine the risk factors associated with Microvascular complications of DM.

Among the 314 patients with DM, 58% were women. The overall prevalence of Microvascular complications (MVCs) is 36.62%. Diabetic peripheral neuropathy was the most frequent complication (27.4%), followed by retinopathy (10.8%) and nephropathy (8%). Inadequate physical exercise was a risk factor for all MVCs. Age, marital status, and level of education were risk factors for neuropathy while smoking and alcohol intake were risk factors for nephropathy. Additionally marital status was a risk factor for retinopathy. Further, the analysis revealed the risk of non-smokers getting neuropathy reduced by 98% compared to smokers (P-0.0001; CI 0.003058-0.191502). lack of regular physical exercise was significantly associated with retinopathy (P 0.012; CI 0.046816-0.690638).

One in ten patients with DM had a microvascular complication. The benefits of microvascular complications prevention should thus be factored into the management of patients with diabetes mellitus.

## Introduction

Diabetes mellitus is a growing global epidemic that disproportionately affects economic and health outcomes in low- and middle-income countries (LMICS) (1). DM is the most common type of diabetes, accounting for around 90% of all diabetes worldwide (2) Diabetic complications are usually divided into two main categories, Macrovascular, and Microvascular complications. Microvascular complications include diabetes retinopathy, nephropathy, and neuropathy (3).

Microvascular complications such as Diabetes retinopathy, nephropathy and peripheral neuropathy are most common complications in diabetes. The global prevalence of this Microvascular complications varies considerably. This is attributable to the population characteristics, screening and treatment methods used in different regions (4).

Diabetes peripheral neuropathy is the most common Microvascular complication across all the regions. Globally the prevalence it is estimated to be18.8%, while Africa has a prevalence of 14.5% (5). The prevalence of DR is estimated to be 35%; out of this 12% are at risk of losing eye sight. Likewise, between 10% and 67% of patients with different levels of renal failure and renal disease is attributed to diabetes. More than 80% of end-stage renal disease (ESRD) is caused by diabetes (2).

The African Region has the highest predicted increase in the number of diabetes cases by 48% and 143% in year 2030 and 2045 respectively. In 2019, 6.8% of mortality was attributable to diabetes with majority of deaths reported in people below 60 years of age. Of the total number of deaths attributable to diabetes, large proportions occur in low- and middle-income countries (41.8% and 58.2%, respectively. It is also estimated (3.9%) 19.4 million adults aged 20–79 years have diabetes. However, more than half (59.7%) of the people living with diabetes are undiagnosed and this could be the reason of low prevalence in the region. Out of 19.4 million reported diabetes cases 8.8% are aged between 65-69 years. Moreover, 5.9% and 2.4% live in urban and rural areas respectively (2).

About 2.4% of Kenyan population are diabetic, 3.1% are pre-diabetic and a substantial proportion of 52.8% people with undiagnosed diabetes are at risk of developing complications. Only 21.3 % of the patients with diabetes reported to be on treatment. Glycemic control was achieved in 7% of the patients on treatment (6). This low glycemic control levels could result to development of complications and premature deaths. The burden of non-communicable diseases continues to increase in Nyeri County having a diabetes prevalence of 6.4% which is almost triple of national prevalence. Almost a half of deaths are due to NCDs i.e. two out of three deaths. Besides, the prevalence of CKD stages 3 to 5 in this adult population who are above 30 years, with type 2 diabetes, is 39.0% (7). As the prevalence of diabetes continues to rise and the burden of diabetic Microvascular complications will increase in future, People with diabetes have an increased risk of developing Microvascular complications, which, if undetected or left untreated, can have a devastating impact on quality of life and place a significant burden on health care costs (8).

Therefore, this study aims to assess the prevalence of diabetes Microvascular complications and associated risk factors among diabetes mellitus patients attending Nyeri County Referral Hospital, Kenya.

## Methodology

### Study design and setting

A hospital-based cross-sectional study was conducted from 08^th^ August 2022 to 31^st^ October 2022 in Nyeri County Referral Hospital in the Diabetic Clinic and Diabetes Outpatient Clinic (DOPC). The diabetes clinic runs every day except weekends and public holiday while the DOPC only runs only of Friday. Both clinics provides all essential general healthcare services to DM patients.

### Study participants

The study recruited diabetes patients aged at least 18 years on follow-up and treatment for at least six months in Nyeri County Referral Hospital in the Diabetic Clinic or Diabetes Outpatient Clinic (DOPC). Participants with a complete follow-up and management documentation in the file were included. participants excluded from the study include those with gestational diabetes, those unstable and required urgent referral and evidence of impaired cognitive function. All the Participants provided informed consent prior to their inclusion in the study.

### Data Sources and measurements

Data was collected from all eligible and consenting participants using a validated researcher administered semi structured questionnaire to collect data on socio-demographic, characteristics and various risk factors. The information collected from the subjects mainly included socio-demographic characteristics (such as age, gender, marital status, education, monthly income, social security, locality type, and ethnicity) and bio-clinical information (such as disease duration, treatment, physical activity, systolic and diastolic blood pressure, waist circumference, body mass index (BMI), biochemical parameters, and presence of complications). Standard and validated protocols were used to classify various biochemical parameters. Details of co-morbidities were obtained from their records as well as from history. Further all the participants were examined for presence of neuropathy, retinopathy and neuropathy.

### Sample size

sample size was calculated using the Cochran’s formula for single population proportion. The prevalence of reported neuropathy, retinopathy and nephropathy in other areas in Kenya is 41, 33 and 0.9% respectively. Based on this finding from previous studies 41% prevalence rate (p) of diabetic neuropathy complication was used in margin of error (d 0.05); and 95% confidence interval. Since the target population was below 10,000 a finite correction formula was applied to get a working sample size of 314 participants. Systemic random sampling was used to collect data from the patients; where the first subject was selected randomly and then every sixth subject was selected for the study till sample size was achieved.

### Data analysis

Data collected was entered and analyzed using STATA Version 17. Descriptive statistics such as mean, frequency, percentages, means and standard deviation was computed for continuous and categorical variables. Chi-square test was used in analysis of categorical variables to test for associations between independent and dependent variables of the subjects. Significant variables from Univariate analysis were analyzed further using binary logistic regression and binomial logistic regression to determine the association of independent risk factors with diabetes Microvascular complications. Results were expressed as odds ratios (OR) and 95% confidence intervals (CI). A p-value of < 5% is considered statistically significant.

### Ethics statement

Both written and verbal informed consent for participation was also obtained from all the participants. For written consent all participants signed the consent form in the questionnaire. Verbal consent was witnessed by the nurse or clinical officer in-charge of the clinic. The study was approved by the Ethical Review Committee of Jomo Kenyatta University of Agriculture and Technology, the National Commission for Science, Technology and Innovation (NACOSTI/P/22/18990) and the County Director of Health Services in Nyeri County.

## Results

### Socio-demographic characteristics of the study participants

Among 314 T2DM patients who participated in this study, 58 % were females, and 42% were males. The participants’ average age was 58.49. ± 17.43 years and 66.6% were married. More than half of the subjects (90.5%) had attained primary education. Of all the subjects in the study, 79% were rural residents (Table 1).

**Table 1:**
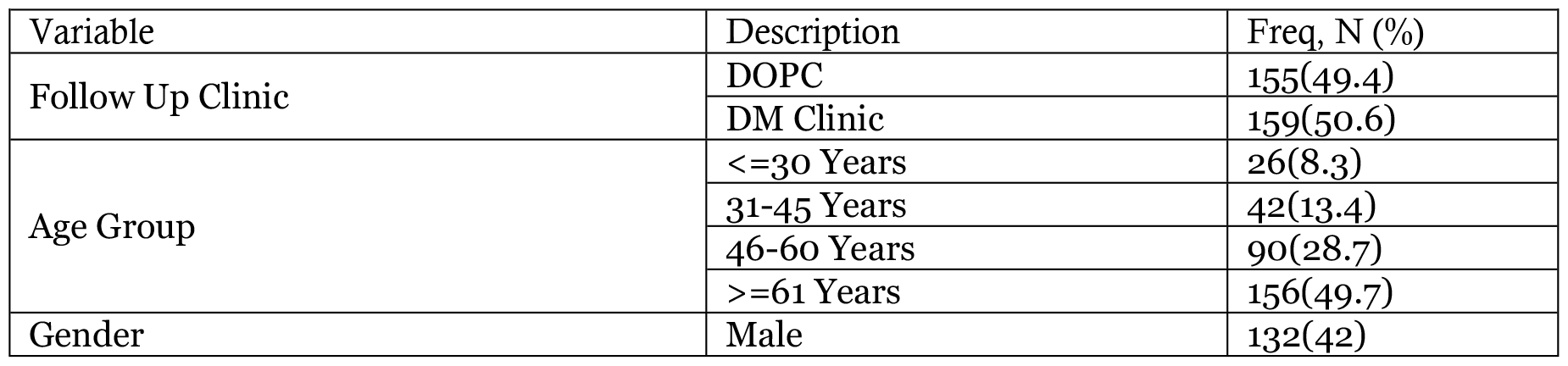
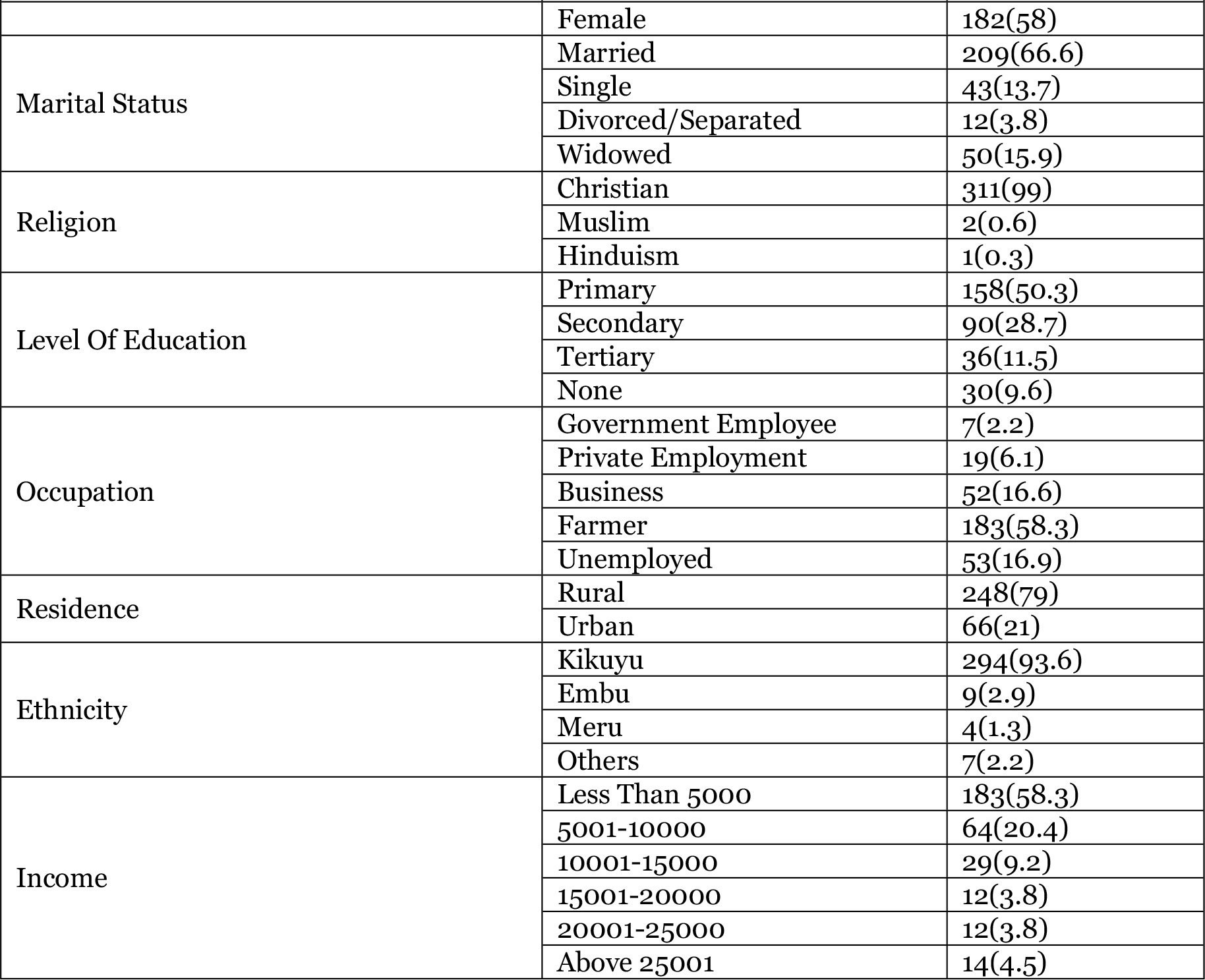
socio demographic characteristics of patients with MVCs.

### Prevalence of Diabetes Microvascular complications and distribution of prevalence by demographic characteristics

The overall prevalence of microvascular complications (MVCs) is 36.62%. Peripheral neuropathy was the most common complication affecting 86(27.4%) of the participants; while retinopathy was at 34(10.8%) and nephropathy was the least 25(8%). Majority of the respondents with diabetic neuropathy complications were from the DOPC 49(31.6%) and more common among those aged >=61 years 56(35.9%). More than half 16 (53.3%) of the respondents with diabetic neuropathy had no formal education.

Diabetic nephropathy complication majorly affected 14(9%) of individuals from the DOPC clinic. It was most common among those aged >=61 years 18(11.5%) and males were more affected 14(10.6%). Additionally, respondents with no formal education and those with rural residence were more affected at 5(16.7%) and 22(8.9%) respectively. DOPC clinic recorded more patients 23(14.8%) with diabetic retinopathy. Retinopathy complication was most common among those aged >=61 years 21(13.5.%) and least common 1(4%) amongst those aged <=30 years. Diabetic retinopathy was most prevalent 4(21.1%) among those in private employment compared to government employees who reported zero prevalence (Table 2).

**Table 2:** Prevalence of Diabetes Microvascular complications and distribution of prevalence by demographic characteristics.

|  |  |  |  |  |  |
| --- | --- | --- | --- | --- | --- |
| Characteristic Variable | Total (N) |  | Overall prevalence (%) |  |  |
|  | 314 |  | 36.62 |  |  |
|  |  |  | Neuropathy (%) | Nephropathy (%) | Retinopathy (%) |
| Microvascular complications | 314 |  | 86 (27.4%) | 25 (8.0%) | 34 (10.8%) |
|  |  | Total participants | Distribution of Prevalence by Demographic characteristics |  |  |
| Follow Up Clinic | DOPC | 155 | 49(31.6%) | 14(9%) | 23(14.8%) |
|  | DM Clinic | 159 | 37(23.3%) | 11(6.9%) | 11(6.9%) |
| Age Group | $\leq 30$ Years | 26 | 1(4%) | 2(8%) | 1(4%) |
|  | 31-45 Years | 42 | 8(19%) | 3(7.1%) | 4(9.5%) |
|  | 46-60 Years | 90 | 21(23.3%) | 2(2.2%) | 8(8.9%) |
| | $\geq 61$ Years | 156 | 56(35.9%) | 18(11.5%) | 21(13.5%) |
| Gender | Male | 132 | 33(25%) | 14(10.6%) | 10(7.6%) |
|  | Female | 182 | 53(29.1%) | 11(6%) | 24(13.2%) |
| Marital Status | Married | 209 | 57(27.3%) | 14(6.7%) | 23(11%) |
|  | Single | 43 | 5(11.6%) | 2(4.7%) | 1(2.3%) |
|  | Divorced/Separated | 12 | 1(10%) | 1(10%) | 0(0%) |
|  | Widowed | 50 | 23(46%) | 8(16%) | 10(20%) |
| Religion | Christian | 311 | 85(27.3%) | 25(8%) | 34(10.9%) |
|  | Muslim | 2 | 1(50%) | 0(0%) | 0(0%) |
|  | Hinduism | 1 | 0(0%) | 0(0%) | 0(0%) |
| Education | Primary | 158 | 42(26.6%) | 15(9.5%) | 16(10.1%) |
|  | Secondary | 90 | 21(23.3%) | 3(3.3%) | 6(6.7%) |
|  | Tertiary | 36 | 7(19.4%) | 2(5.6%) | 6(16.7%) |
|  | None | 30 | 16(53.3%) | 5(16.7%) | 6(20%) |
| Occupation | Government Employee | 7 | 0(0%) | 0(0%) | 0(0%) |
|  | Private Employment | 19 | 7(36.8%) | 0(0%) | 4(21.1%) |
|  | Business | 52 | 15(28.8%) | 5(9.6%) | 4(7.7%) |
|  | Farmer | 183 | 50(27.3%) | 12(6.6%) | 19(10.4%) |
|  | Unemployed | 53 | 14(26.4%) | 8(15.1%) | 7(13.2%) |
| Residence | Rural | 248 | 71(28.6%) | 22(8.9%) | 27(10.9%) |
|  | Urban | 66 | 15(22.7%) | 3(4.5%) | 7(10.6%) |

### Risk factors associated with diabetes retinopathy, nephropathy and neuropathy

physical exercise, age cohort, marital status and level of education had a significant relationship with neuropathy complications were with p values of 0.002, 0.001, 0.002 and 0.007 respectively. A significant relationship exists between nephropathy complications and smoking, history of alcohol intake, physical exercise and HBIAC test with p values of 0.001, 0.001, 0.0002 and 0.011 respectively.002, 0.001, 0.002 and 0.007 respectively. Additionally, a significant relationship with retinopathy complications were physical exercise and marital status with p values of 0.013 and 0.033 respectively (Table 3).

**Table 3:** Risk factors associated with diabetes retinopathy, nephropathy and neuropathy among diabetes mellitus patients.

|  |  |  |  |  |  |  |  |
| --- | --- | --- | --- | --- | --- | --- | --- |
| Variable | Variable choices | Type of Complication and P Value |  |  |  |  |  |
|  |  | Neuropathy Yes, n (%) | P value | Nephropathy Yes, n (%) | P value | Retinopathy Yes, n (%) | P value |
| BMI | Normal | 37(29.6) | 0.236 | 10(%yes) | 0.633 | 15(12) | 0.864 |
|  | Under weight | 0(0) |  | 0(0) |  | 0(0) |  |
|  | Over weight | 40(29.9) |  | 13(9.7) |  | 15(11.2) |  |
|  | Obese | 9(17.6) |  | 2(3.9) |  | 4(7.8) |  |
| Smoke | Yes | 3(17.6) | 0.267 | 5(29.4) | 0.001 | 0(0) | 0.233 |
|  | No | 83(27.9) |  | 20(6.7) |  | 34(11.4) |  |
| history of alcohol intake | Yes | 4(23.5) | 0.481 | 5(29.4) | 0.001 | 1(5.9) | 0.705 |
|  | No | 82(27.6) |  | 20(6.7) |  | 33(11.1) |  |
| physical exercise | Yes | 62(23.8) | 0.002 | 15(5.8) | 0.002 | 23(8.8) | 0.013 |
|  | No | 24(44.4) |  | 10(18.5) |  | 11(20.4) |  |
| age group | <=30 years | 1(3.8) | 0.001 | 2(7.7) | 0.078 | 1(3.8) | 0.494 |
|  | 31-45 years | 8(19) |  | 3(7.1) |  | 4(9.5) |  |
|  | 46-60 years | 21(23.3) |  | 2(2.2) |  | 8(8.9) |  |
|  | >=61 years | 56(35.9) |  | 18(11.5) |  | 21(13.5) |  |
| Gender | Male | 33(25) | 0.249 | 14(10.6) | 0.14 | 10(7.6) | 0.114 |
|  | Female | 53(29.1) |  | 11(6) |  | 24(13.2) |  |
| marital status | Married | 57(27.3) | 0.002 | 14(6.7) | 0.142 | 23(11) | 0.033 |
|  | Single | 5(11.6) |  | 2(4.7) |  | 1(2.3) |  |
|  | Divorced/separated | 1(8.3) |  | 1(10) |  | 0(0) |  |
|  | Widowed | 23(46) |  | 8(16) |  | 10(20) |  |
| level of education | Primary | 42(26.6) | 0.007 | 15(9.5) | 0.078 | 16(10.1) | 0.134 |
|  | secondary | 21(23.3) |  | 3(3.3) |  | 6(6.7) |  |
|  | Tertiary | 7(19.4) |  | 2(5.6) |  | 6(16.7) |  |
|  | None | 16(53.3) |  | 5(16.7) |  | 6(20) |  |
| employment status | government employee | 0(0) | 0.497 | 0(0) | 0.206 | 0(0) | 0.482 |
|  | Private | 7(36.8) |  | 0(0) |  | 4(21.1) |  |
|  | Business | 15(28.8) |  | 5(9.6) |  | 4(7.7) |  |
|  | Farmer | 50(27.3) |  | 12(6.6) |  | 19(10.4) |  |
|  | Unemployed | 14(26.4) |  | 8(15.1) |  | 7(13.2) |  |
| Variable | Variable choices | Type of Complication and P Value |  |  |  |  |  |
|  |  | Neuropathy Yes, n (%) | P value | Nephropathy Yes, n (%) | P value | Retinopathy Yes, n (%) | P value |
| current treatment | Metformin 500mg | 17(24.6) | 0.784 | 2(2.9) | 0.191 | 12(17.4) | 0.141 |
|  | Metformin 500mg and Glibenclamide | 8(28.6) |  | 2(7.1) |  | 1(3.6) |  |
|  | Metformin 850/1000mg | 28(29.5) |  | 7(7.4) |  | 11(11.6) |  |
|  | Metformin+Glibenclamide+Insulin | 20(31.7) |  | 5(7.9) |  | 8(12.7) |  |
|  | Insulin only | 13(22.8) |  | 9(15.8) |  | 2(3.5) |  |
|  | Others | 0(0) |  | 0(0) |  | 0(0) |  |
| HBIAC Test | Normal | 37(29.6) | 0.475 | 4(3.2) | 0.011 | 17(13.6) | 0.199 |
|  | Abnormal | 49(25.9) |  | 21(11.1) |  | 17(9) |  |

### Regression analysis for risk factors for diabetes Microvascular complications

Controlling for frequency of exercise, marital status and level of education; age cohorts were not significantly associated with neuropathic complications. Similarly holding age cohorts, frequency of exercise and level of education constant, marital status were not significantly associated with neuropathic complications (Table 4).

**Table 4:** Regression analysis for risk factors for diabetic neuropathy.

| Table 4: Regression analysis for risk factors for diabetic neuropathy |  |  |  |  |  |  |
| --- | --- | --- | --- | --- | --- | --- |
| Complication Neuropathy | Odds Ratio | Std. Err. | Z | P>z | [95% Conf. Interval] |  |
| Age cohorts (Base-<=30years) |  |  |  |  |  |  |
| <=30 years | 1 | (base) |  |  |  |  |
| 31-45 years | 0.452414 | 0.243326 | -1.47 | 0.14 | 0.157662 | 1.298211 |
| 46-60 years | 0.730886 | 0.26773 | -0.86 | 0.392 | 0.35649 | 1.498484 |
| >=61 years | 1 | (omitted) |  |  |  |  |
| marital status (Base-Married) |  |  |  |  |  |  |
| Single | 0.832651 | 0.509795 | -0.3 | 0.765 | 0.25079 | 2.764494 |
| Divorced/Separated | 0.376244 | 0.410275 | -0.9 | 0.37 | 0.044389 | 3.189042 |
| Widowed | 1.489481 | 0.647433 | 0.92 | 0.359 | 0.635394 | 3.49162 |
| level of education (Base-Primary) |  |  |  |  |  |  |
| Secondary | 1.383951 | 0.517554 | 0.87 | 0.385 | 0.664965 | 2.880334 |
| Tertiary | 1.182695 | 0.65043 | 0.31 | 0.76 | 0.402485 | 3.475331 |
| None | 1.893705 | 1.055392 | 1.15 | 0.252 | 0.63522 | 5.645478 |
| Frequency of exercise (Base-Daily) |  |  |  |  |  |  |
| Once weekly | 0.811892 | 0.360899 | -0.47 | 0.639 | 0.339724 | 1.940308 |
| 2-3 times a week | 0.536864 | 0.204653 | -1.63 | 0.103 | 0.254323 | 1.133295 |
| 2 weeks to one month | 1 | (empty) |  |  |  |  |
| Only when recommended by HC person | 0.767539 | 0.928569 | -0.22 | 0.827 | 0.071667 | 8.220229 |
| _cons | 0.522596 | 0.205271 | -1.65 | 0.099 | 0.242003 | 1.128524 |

Holding for age cohorts, frequency of exercise and level of education, the odds of those who are not smoking getting nephropathy reduces by 98% compared to smokers (P 0001; CI 0.003058-0.191502) (Table 5).

**Table 5:** Regression analysis for risk factors for diabetic nephropathy.

| Complication Nephropathy | Odds Ratio | Std. Err. | Z | P>z | [95% Conf. Interval] |  |
| --- | --- | --- | --- | --- | --- | --- |
| Age cohorts (Base-<=30years) |  |  |  |  |  |  |
| <=30 | 1 | (base) |  |  |  |  |
| 31-45 | 1 | (empty) |  |  |  |  |
| 46-60 | 0.22572 | 0.430797 | -0.78 | 0.435 | 0.005358 | 9.508858 |
| >=61 | 4.504898 | 7.633377 | 0.89 | 0.374 | 0.162688 | 124.7422 |
| Frequency of exercise (Base-Daily) |  |  |  |  |  |  |
| Once weekly | 4.175531 | 4.012703 | 1.49 | 0.137 | 0.634897 | 27.46123 |
| 2-3 times a week | 0.68741 | 0.649369 | -0.4 | 0.692 | 0.107924 | 4.378367 |
| 2 weeks to one month | 1 | (empty) |  |  |  |  |
| Only when recommended by HC person | 1 | (empty) |  |  |  |  |
| level of education (Base-Primary) |  |  |  |  |  |  |
| Secondary | 0.277528 | 0.328682 | -1.08 | 0.279 | 0.02724 | 2.827485 |
| Tertiary | 1.612873 | 1.707157 | 0.45 | 0.652 | 0.2026 | 12.83987 |
| None | 0.568066 | 0.574989 | -0.56 | 0.576 | 0.078131 | 4.130236 |
| Smoking (Base-Yes) |  |  |  |  |  |  |
| No | 0.024201 | 0.025541 | -3.53 | 0.0001 | 0.003058 | 0.191502 |
| hb1ac | 2.217372 | 1.664154 | 1.06 | 0.289 | 0.509338 | 9.653207 |
| _cons | 0.38589 | 0.624194 | -0.59 | 0.556 | 0.016204 | 9.190042 |

Adjusting for marital status, the odds of those who do exercise once weekly getting retinopathy complication reduces by 83% compared to those who do exercise daily (P 0.012; CI 0.046816-0.690638). Controlling for marital status, the odds of those who do exercise 2-3 times a week reduces by 87% compared to those who do exercise daily (P 0.0001; CI 0.049133-0.397895) (Table 6).

**Table 6:**
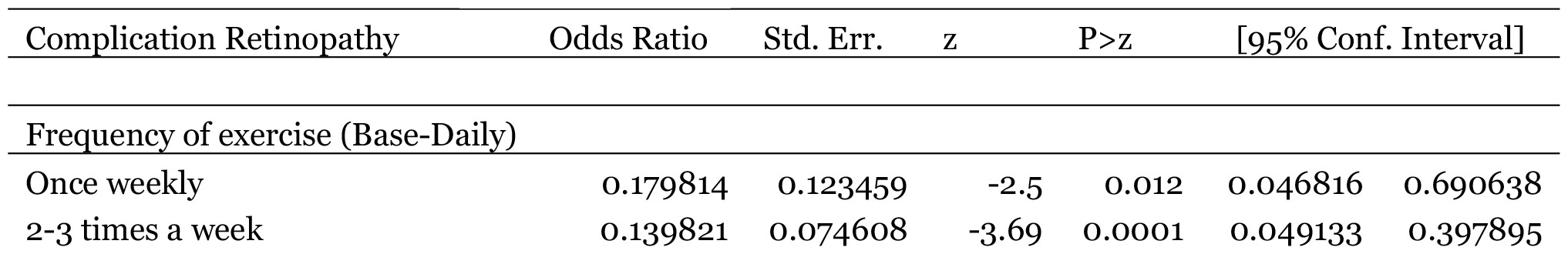
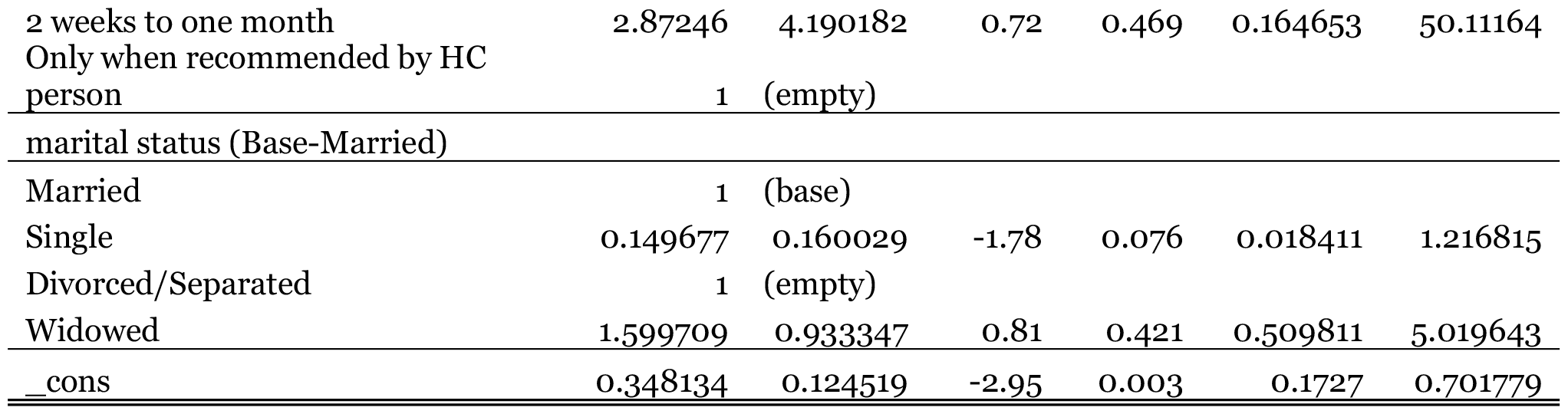
Regression analysis for risk factors for diabetic Retinopathy.

## Discussion

The current study sought to assess the prevalence of and the associated risk factors among patients attending Nyeri County Referral Hospital in Kenya. The study observed that overall prevalence of microvascular complications is 36.62%. Among the microvascular complications, peripheral neuropathy had the highest prevalence (27.4%) followed by retinopathy (10.8%) and nephropathy (8 %). These findings are similar to findings from studies other studies conducted in other low income countries such as Ethiopia, Tanzania and India (9–11).However, this findings might differ in specific percentage of proportions the difference might be related to accessibility and advancement of health institutions and patient’s adherence to medication and practice in different regions.

Univariate analysis showed that factors such as lower education, occupation i.e. farmers and patient who resided in the rural areas were significantly associated with peripheral neuropathy (P < .05). Lower level of education, unemployment, smoking and patients who resided in the rural areas were significantly associated with nephropathy (P < .05) whereas lower education, frequency of exercise and private employment were significantly associated with retinopathy (P < .05). All the three MVCs prevalence was higher in patients seen in the Diabetes outpatient clinic (DOPC) when compared to patients seen in the diabetes clinic. The high prevalence in DOPC could be due to delay in diagnosis, poor self-care, poor health-seeking behavior as the clinic runs only once per week and patients are given appointment of up to 3 months while the other clinic runs daily and patients are likely are given a nearer return date. Lower education status was an independent and commonly encountered risk factors for all the MVCs. Educational status influences the awareness about diabetes, compliance to drugs and the health seeking behavior of an individual. Studies have observed that lower the education higher is the risk of developing diabetes complications. Agriculture is the major occupation in rural areas in Kenya. Farmers are more prone to foot injuries during work this could be reason for high prevalence of neuropathy (10,12).

The presence of DPN and Nephropathy was associated with rural sector of residence and lower household income. A possible explanation for the occurrence could be that poor people are less likely to use health services, which might result in delayed diagnosis and poor control of DM. Previous studies have reported that metabolic control of DM was worse in patients with a lower socio-economic status. In addition, the increased risk in rural sector residents could be due to lack of access to the better health care facilities available to residents in urban areas (13).

Multi-regression analysis revealed that the risk of diabetic Retinopathy reduces by 87% in those who do exercise 2-3 times a week compared to those who do exercise daily (P 0.0001; CI 0.049133-0.397895. Even though there is strong evidence for the correlation between physical activity and disease status, the number of people who actually partake in the minimum requirement for physical activity as prescribed by the American College of Sports of Medicine (ACSM) and other Public Health and Exercise Authorities remains low around the world. Physical activity (PA) is a critical component of lifestyle intervention in diabetes management. This study agrees with other studies which revealed that total physical activity was decreased in patients with severe to very severe non-proliferative and proliferative diabetic retinopathy (14,15) Adjusting for age cohorts, frequency of exercise and level of education, the odds of those who are not smoking getting nephropathy reduces by 98% compared to smokers (P-0.0001; CI 0.002549-0.145117). Smoking, a well-known and preventable risk factor for many diseases. Cigarette smoking in diabetes has been repeatedly confirmed as an independent risk factor for the onset and progression of diabetic nephropathy. This study demonstrates strong association found between chronic cigarette smoking and diabetic nephropathy. This is consistent with various studies that suggest; smoking is a major fuel in the development of high oxidative stress and subsequently hyperlipidemia, accumulation of advanced glycation end products, activation of the renin angiotensin system and Rho-kinase, which are observed to play a pathogenic role in the progression of diabetic nephropathy. Furthermore, cigarette smoking in diabetic patients with vascular complications produces a variety of pathological changes in the kidney, such as thickening of the glomerular basement membrane and mesangial expansion with progression in glomerulus sclerosis and interstitial fibrosis, which ultimately results in end stage renal failure (16,17)

### Study Limitations

This study is a cross-sectional study hence cannot establish causal inferences but rather associations. Interpretation of findings should therefore be done cautiously. Nonetheless, this study provides some estimates on the prevalence of MVCs and associated risk factors among DM patients inform development of interventions. The sample size of 314 is also large enough to draw valid conclusions from a single center study.

## Conclusion

In this study, four in ten patients with DM had a microvascular complication. Three in ten had a neuropathy and two in ten had a retinopathy. Factors such as physical activity, dietary practice, level of education, clinic where followed-up and area of residence being associated with the MVCs. Identifying patients at risk of MVCs can lead to targeted interventions in line with the Kenyan government NCD policy and SDG 3.4 target on reducing premature mortality from NCDs through prevention and treatment. Additionally, public health efforts should institute measures to ensure that health promotion and education on long-term diseases like DM form part of the routine management and care practices.

## Data Availability

All data is included in tables

## Acknowledgments

The authors would like to thank the study facilities and participants for their role in this study as well as clinicians in the DOPC and DM clinic for their support

## Authors’ contribution

Rachael Ireri conceptualized the study, designed the study, collected and analysed data, interpreted data, prepared and the reviewed manuscript for the final submission. Gideon Kikuvi, Susan Mambo and Besty Cheriro formulated research question and assisted and validated the manuscript for the final submission

